# Automated language impairment screening in acute stroke using connected speech

**DOI:** 10.64898/2026.08.14.26360474

**Authors:** Lokesha S. Pugalenthi, Tatiana T. Schnur

## Abstract

Connected speech is essential for everyday communication, but clinical constraints and patient fatigue limit detailed evaluation in acute stroke (<1-week post-stroke). Bedside assessments may sample discourse but rarely quantify language impairment (LI) in connected speech, leaving patients’ communication poorly characterized. We analyzed brief story retellings from 86 patients with left-hemisphere stroke (∼4 days post-stroke; 63 classified with LI using composite clinical and naming criteria). From transcripts generated with automatic speech recognition, we derived discrete linguistic features and embeddings with Large Language Models (LLMs). An ensemble of embedding-based classifiers distinguished patients with and without LI with 90% balanced accuracy (79% sensitivity, 100% specificity), outperforming independent embedding and discrete-linguistic-based classifiers, showing distinct LLMs contributed complementary information. Adding the discrete-linguistic-based classifier to the ensemble did not improve balanced accuracy but modestly increased sensitivity at the expense of specificity. We provide proof of concept for a fast, largely automated discourse screener of acute LI.

---

While a stroke in the left hemisphere disrupts language at multiple levels, including retrieving single words and producing grammatical sentences, the ability to produce coherent, connected speech (i.e., discourse) is arguably the most fundamental requirement for real-world human communication^1,2^. However, detailed assessments of connected speech are rarely conducted at the acute stage of stroke (<1-week post-stroke onset)^3,4^ because acute patients are often medically unstable and fatigued, and clinicians must prioritize evaluating immediate needs like swallowing function (dysphagia). Although, standard bedside evaluations like the National Institute of Health Stroke Scale^5^ typically sample discourse providing an established clinical benchmark for gross language status, their language evaluation relies on manual, subjective clinician scoring which does not quantify subtle or complex language impairments in discourse. Compounding these challenges, existing assessments of discourse can be insufficiently validated for acute settings^4,6,7^. Thus, acute language impairment affecting complex, real-world narrative production remains largely unquantified, despite its importance for everyday functioning and independence.

Story retelling offers a brief, feasible, and ecologically valid alternative for capturing naturalistic connected speech at bedside. Unlike static picture descriptions, which often elicit isolated lists of nouns and simple action verbs, story retellings require narrative coherence and temporal sequencing, key features of real-world discourse^8,9^. Furthermore, because story retellings yield representative discourse samples in just 1–2 minutes, they can be readily acquired from acute patients with remaining verbal capacity without inducing over-fatigue or overtaxing clinicians ^7,8,10-12^.

In a bid to capture discourse impairments without overextending patients or clinicians, previous studies applied automatic speech recognition (ASR) and natural language processing (NLP) to discourse recordings to derive discrete linguistic features as input to machine learning (ML)-based classification algorithms. This approach achieved near-perfect accuracy in differentiating people with language impairment at the chronic stage of stroke (>6 months post-stroke onset) from neurologically healthy controls^13-15^. However, this approach relies on extracting lists of discrete linguistic features, such as informativeness and the number of accurate and relevant words, which may not provide a holistic representation of a discourse sample. Further, in the acute setting, relevant work remains limited to group-level analyses of manually transcribed picture descriptions^16,17^. This limits immediate clinical applicability and ecological validity, as manual transcription is labor-intensive and picture descriptions are less sensitive to real-world communicative abilities compared to story retelling^8,9^. Moreover, the primary clinical and scientific challenge is not distinguishing impaired individuals from neurologically healthy controls, but differentiating acute post-stroke individuals with versus without language impairment, who may present with subtle discourse impairments^18^.

Large Language Models (LLMs) offer a powerful, automated computational alternative to discrete linguistic feature extraction. LLMs contextually evaluate connected language by encoding the holistic meaning of a discourse sample into a single high-dimensional vector (i.e., an embedding), where semantically similar samples (e.g., “Cinderella is a girl,” “Cinderella is a woman”) yield similar embeddings^18^. An acknowledged limitation of embedding-based classifiers is their reduced interpretability relative to discrete linguistic features, as the specific dimension driving classification cannot be easily defined. Nevertheless, in neurodegenerative patient cohorts, LLM-derived embeddings from manual transcriptions of discourse samples successfully distinguished patients with neurodegenerative cognitive impairment from neurologically healthy controls (>77% accuracy)^18,19^, demonstrating that high-dimensional embeddings capture discourse properties critical for language breakdown. However, whether LLM embeddings accurately identify acute language impairment from ASR-transcribed discourse samples of naturalistic post-stroke speech, and whether high-dimensional embeddings outperform traditional, interpretable discrete linguistic features, remains an open question.

Furthermore, previous language classification research evaluated individual LLMs, leaving the potential of an ensemble model consisting of multiple LLMs unexplored. Although ensemble modeling generally boosts predictive accuracy by leveraging predictions across multiple classifiers^20^, whether this approach improves language impairment classification from naturalistic speech remains unknown. Because different LLMs are trained on distinct corpora with varying pre-training objectives, an ensemble model of embedding-based classifiers across diverse architectures may capture complementary aspects of impaired discourse, ranging from local semantic density to global narrative coherence, which may enhance classification performance.

To address these gaps, the goal of the current study was to develop a brief, ecologically valid, and largely automated digital screener capable of accurately identifying language impairment in acute post-stroke individuals from a brief narrative. In contrast to previous acute post-stroke discourse studies that rely on labor-intensive transcription and hand-coded metrics, our framework leverages ASR for automated transcript generation of preprocessed audio, NLP for discrete feature extraction, and derives embeddings using GloVe^21^ and three distinct LLM architectures (BERT, Mistal, and OpenAI)^22-24^. Across other clinical populations, both discrete linguistic features and embeddings independently show success in capturing language deficits^13-15,18,19^.

The study had three aims: First, to evaluate whether automatically extracted NLP-derived discrete linguistic features successfully classify language impairment in acute post-stroke individuals; second, to test whether high-dimensional LLM embeddings capture language breakdown and outperform traditional discrete linguistic features; and third, to determine whether an ensemble model of embedding classifiers from multiple LLMs, or an ensemble model comprising of the linguistic classifier in addition to the embedding classifiers, provide complementary predictive value. To our knowledge, this study is the first to differentiate, on an individual basis, acute left-hemisphere stroke patients with and without language impairment using largely automated analysis of real-world discourse. Ultimately, by establishing a feasible and largely automated discourse screening pipeline, this approach has the potential to extend acute clinical assessments into the functional domain of connected speech, reduce clinician workloads, and facilitate timely therapeutic referrals to speech therapy during a patient’s initial hospitalization.

## Results

For a summary of overall results, see Figure 1. Confidence intervals are reported in Supplementary Figure 1.

**Figure 1.**
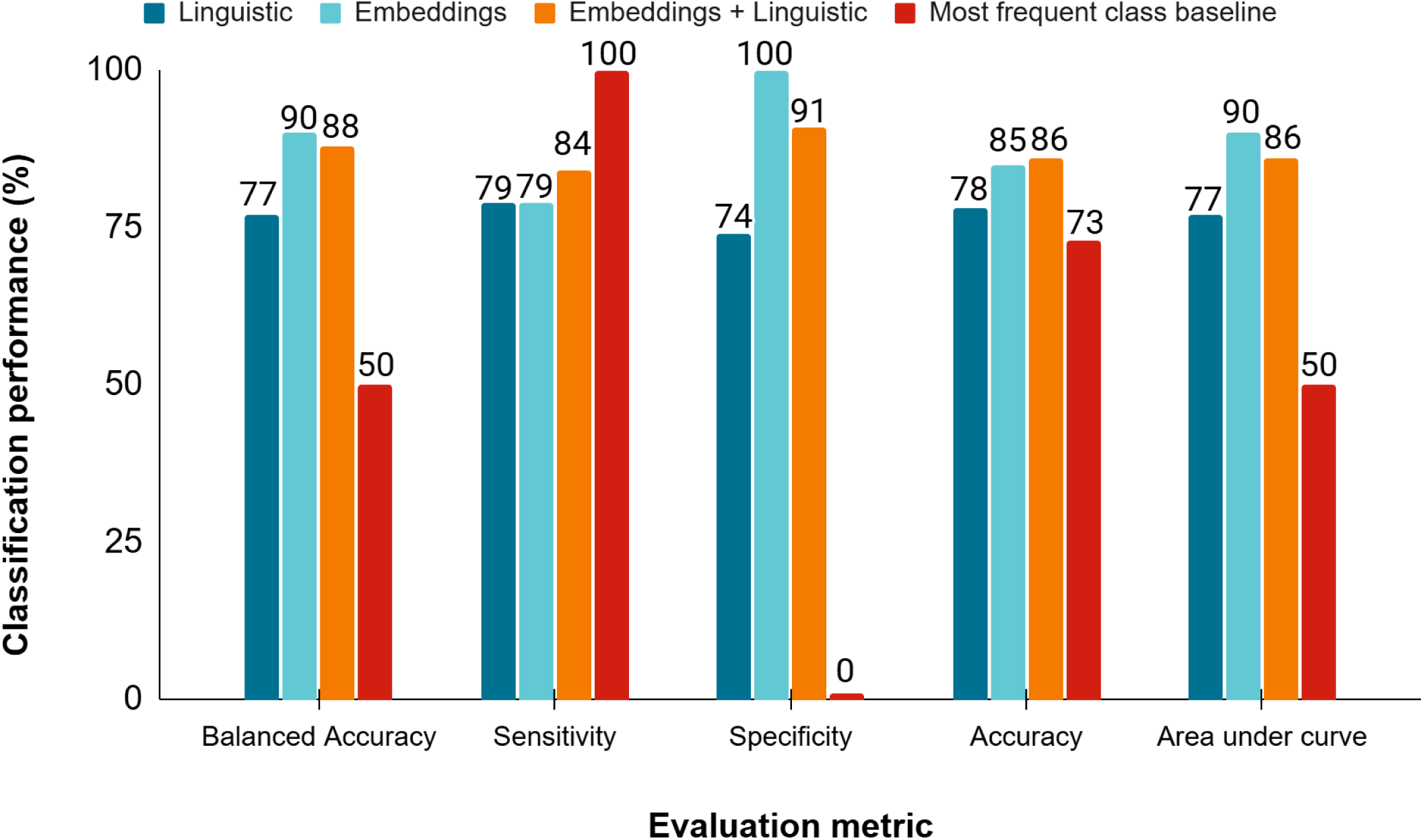
Classification performance of detecting acute post-stroke language impairment from story retellings. Comparison of evaluation metrics (balanced accuracy, sensitivity, specificity, accuracy, and area under the receiver operating characteristic curve (AUC)) for the linguistic classifier, embedding-only ensemble classifier, and the embedding-and-linguistic ensemble classifier. Values are shown as percentages; AUC values multiplied by 100 for display consistency. Sensitivity is the proportion of participants with language impairment correctly identified; specificity is the proportion of participants without language impairment correctly identified. Balanced accuracy is the average of sensitivity and specificity. Accuracy is the proportion of participants correctly identified. Chance-level performance for balanced accuracy and AUC is 50%.

### Linguistic classifier

For the discrete linguistic feature set, among the ML-based classification algorithms evaluated, logistic regression achieved the highest balanced accuracy score (average of sensitivity and specificity; 77%; Figure 1), outperforming the baseline of predicting each sample as the most frequent class in the dataset, namely story retellings from participants identified with acute language impairment, by 27% balanced accuracy. As seen in Figure 2a, the linguistic classifier achieved 79% sensitivity where 50 out of 63 PWA were correctly classified as PWA and 74% specificity, where 17 out of 23 participants without language impairment were correctly classified as participants without language impairment. Supplementary Figure 2 shows the linguistic classifier’s most discriminative features, defined as those causing a ≥2% decrease in classification performance when permuted on average^21^ (see Methods section *Feature importance)*. Six of 14 features met this criterion. See Supplementary Figure 3 for individual values across the discrete linguistic features. See Supplementary Figure 4a for the classification performance of ML-based evaluated classification algorithms: shallow neural network, decision tree, support vector machine, logistic regression, random forest, and gradient boosting.

**Figure 2.**
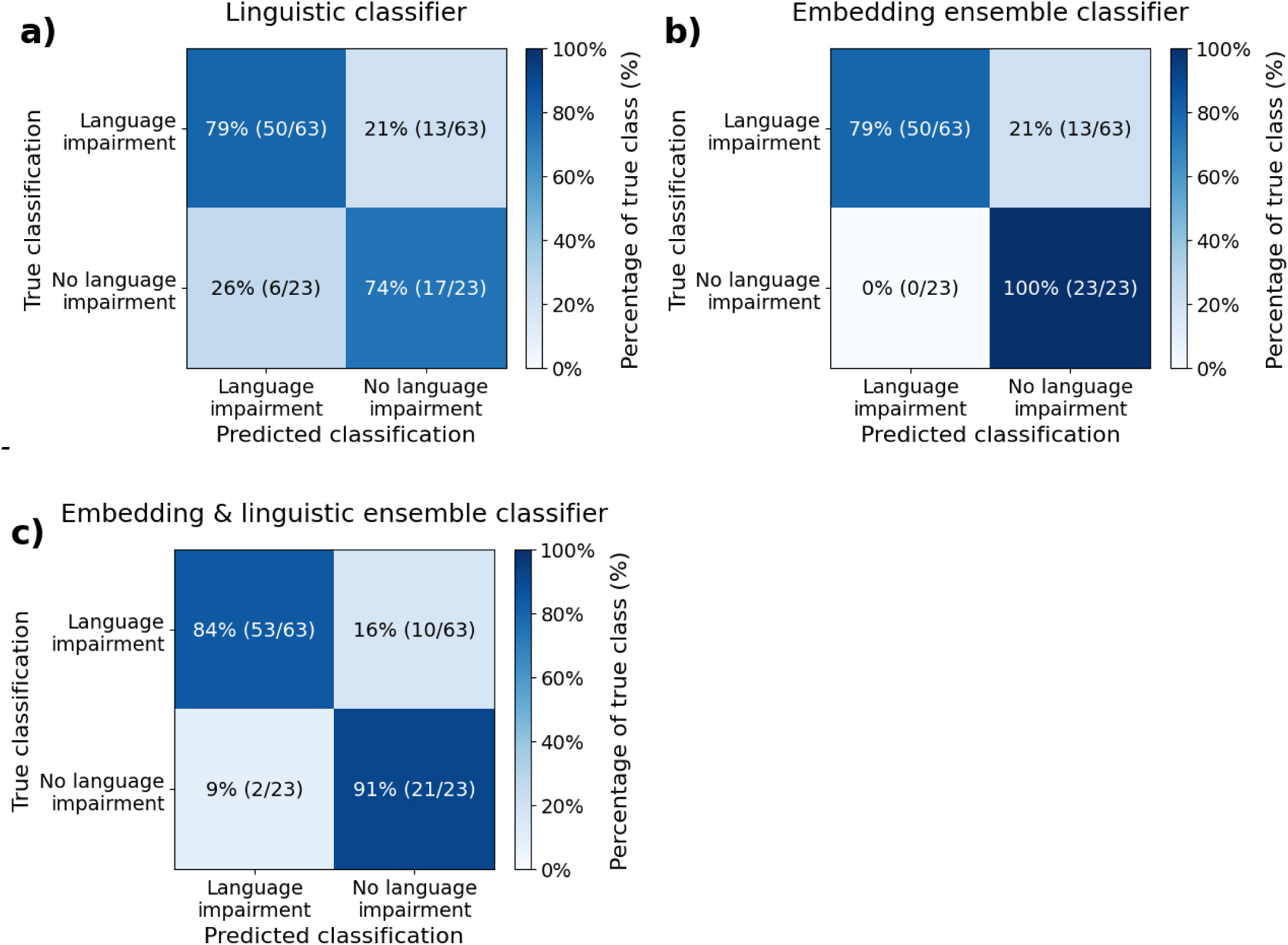
Confusion matrices for the linguistic classifier, embedding ensemble classifier, and embedding-and-linguistic ensemble classifier. Confusion matrices are row-normalized by true classification group. Rows indicate true classification (top: language impairment, n = 63; bottom: no language impairment, n = 23), and columns indicate predicted classification (left: language impairment; right: no language impairment). Because language impairment is the target condition evaluated, the upper-left cell represents sensitivity (true positive rate), and the lower-right cell represents specificity (true negative rate). Cell values display row percentages with raw counts in parentheses; darker shading indicates higher percentages. Diagonal cells represent correct classifications, whereas off-diagonal cells represent misclassifications.

### Embedding-based classifiers

The balanced accuracy of each independent embedding classifier (GloVe^22^, BERT^23^, Mistral^24^, OpenAI^25^) ranged from 72-83% balanced accuracy, with embeddings from LLM Mistral achieving the highest balanced accuracy score (84%, best classification algorithm was support vector machine). See Supplementary Figure 5 for the performance of each ML-based classification algorithm paired with each LLM’s embedding.

The first ensemble model comprising the four embedding-based classifiers achieved a balanced accuracy of 90% (support vector machine, see Figure 1 and Supplementary Figure 4b). As seen in Figure 2b, the embedding-based ensemble classifier achieved 79% sensitivity, where 50 out of 63 PWA were correctly classified as PWA and 100% specificity, where 23 out of 23 participants without language impairment correctly classified as participants without language impairment. The embedding-based ensemble classifier outperformed independent embedding classifiers by ≥6% balanced accuracy. Permutation analysis indicated that when the language impairment prediction probabilities from an LLM’s independent embedding-based classifier were permuted, the classification performance of the embedding-based ensemble classifier dropped on average: OpenAI (16%), Mistral (13%), GloVe (6%), and BERT (3%). The permutation analysis demonstrated that Mistral and Open AI’s embeddings were the most important to the ensemble classifier, while GloVe and BERT’s embeddings were the least important. The embedding-based ensemble classifier outperformed the linguistic classifier by 13% in balanced accuracy (0% increase in sensitivity, 26% increase in specificity). In sum, the ensemble of embedding-based classifiers from multiple LLMs achieved more accurate differentiation than the linguistic classifier and the independent embedding-based classifiers.

### Combining linguistic-based and embedding-based classifiers

The second ensemble model, comprising both the linguistic classifier and the four embedding-based classifiers, achieved a balanced accuracy of 88% (support vector machine, see Figure 1 and Supplementary Figure 4c), performing 2% worse balanced accuracy compared to the embedding-only ensemble model, and 11% better than the linguistic-based classifier. As seen in Figure 2c, this second ensemble model achieved 84% sensitivity where 53 out of 63 PWA were correctly classified as PWA and 91% specificity, where 21 out of 23 participants without language impairment were correctly classified as participants without language impairment. Akin to the embedding-only ensemble classifier, OpenAI and Mistral contributed the most to classification performance (20% and 9% respectively), while GloVe and BERT contributed the least (6% and 3%, respectively). In sum, adding the linguistic classifier to the ensemble model improved only the ensemble’s sensitivity.

## Discussion

In this study, we established a brief digital screener integrating automatic speech recognition (ASR), natural language processing (NLP), and Large Language Model (LLM) embeddings to identify language impairment in acute stroke participants with verbal capacity from 1–2-minute discourse samples^7,8,10-12^. Three main findings emerged. First, brief story retellings acquired at the acute bedside contained sufficient information to support accurate identification of language impairment. Second, an ensemble classifier combining embedding classifiers from multiple LLMs achieved excellent discrimination (90% balanced accuracy) outperforming both the independent embedding classifiers and the traditional linguistic classifier.

Third, adding the linguistic classifier to the embedding ensemble did not improve balanced accuracy (88%), but shifted performance toward greater detection of language impairment, increasing sensitivity from 79% to 84% while reducing specificity from 100% to 91%, a trade-off that may be clinically appropriate when minimizing missed cases. Together, these findings demonstrate that automated analysis of discourse is both feasible and promising for screening language impairment in the acute stroke setting. Below, we discuss how these results advance prior work and outline future directions.

This work advances the analysis of discourse post-stroke in three ways. First, it addressed the clinically relevant problem of differentiating participants with acute stroke who do, versus do not, have language impairment. This is a more challenging and clinically meaningful task than differentiating participants with language impairment post-stroke from neurologically healthy controls, as in previous studies^13-17^. Second, the present approach focused on the individual patient rather than relying on group comparisons of discrete linguistic features between neurologically healthy controls and people with stroke-induced language impairment^16,17^. This individual-level classification more closely matches the decisions clinicians must perform in the acute stroke setting. Third, this classification approach successfully evaluated language in a more real-world context, specifically discourse produced during story retelling^8,9^. By developing an individual-level screener of more real-world communication that accurately predicts the presence or absence of language impairment in the acute stroke clinical setting, we will be able to facilitate referrals for treatments that accelerate language recovery.

The findings also have broader implications for the use of LLM-derived embeddings to automatically detect language impairment from transcribed speech. First, ensembling embedding classifiers from multiple LLMs^22-25^ improved differentiation. With the exception of BERT^23^, permutation analysis demonstrated that each classifier in the embedding-based ensemble classifier contributed unique predictive information (permutation values > 5%). This was also the case for the ensemble classifier that combined both linguistic and embedding-based classifiers. This result implies that different LLMs capture distinct aspects of impaired discourse, suggesting that combining embedding-based classifiers from multiple LLMs may be useful to detect language impairment in other clinical populations as well. To our knowledge, this is the first study to show this in the context of speech-based classification^26^. Among the LLMs we evaluated, Mistral^24^ was the most accurate individually and the second strongest contributor to both ensembles. This is consistent with prior work in chronic stroke that also applied LLMs to speech elicited with Cinderella story retells^15^, suggesting that future studies of discourse post-stroke should prioritize using Mistral-based embeddings. Overall, these findings suggest that future work applying LLM embeddings to post-stroke discourse should consider combining embeddings from multiple LLMs to maximize predictive accuracy and prioritize Mistral-based embeddings.

While these findings establish a proof of concept for automated acute screening, translating this computational pipeline into routine clinical practice presents several opportunities for optimization. First, enhancing end-to-end automaticity will streamline point-of-care deployment. Although we manually trimmed non-participant speech in the current pipeline, integrating automated speaker diarization tools (e.g., Pyannote^27^) can isolate patient speech directly within noisy hospital environments. Resolving this step will enable mobile screening applications where clinicians upload brief audio recordings and receive real-time estimated probabilities of language impairment (c.f. screening apps in other clinical populations with language impairment^28^). Second, addressing the classic trade-off between model accuracy and interpretability^29^ will maximize clinical utility. While LLM embeddings yielded superior classification, their high-dimensional nature offers less transparent feedback than discrete feature counts. Future iterations can bridge this gap by combining LLM fine-tuning (adjusting the weights of an LLM’s final layer to maximize predictive accuracy) with Local Interpretable Model-agnostic Explanations (LIME^30^) methods successfully applied to differentiate subtypes in a primary progressive aphasia cohort^31^. Using LIME, we can highlight specific word-level impairments within a patient’s transcript, providing SLPs with actionable diagnostic targets. Third, predicting continuous impairment severity, rather than relying on binary classification, will improve detection of milder language impairments. Because comprehensive standardized language batteries (e.g., the Western Aphasia Battery^32^) are rarely feasible during acute stroke care, we operationalized ground-truth language impairment status using a triangulated composite criterion from available bedside evaluations. For the 63 participants classified as having language impairment, 40 only met one criterion, suggesting their language impairment was milder than other participants who met multiple criteria (Supplementary Table 1), meaning they are more likely to be misclassified as not having language impairment (i.e. false negative). Nine of our model’s 13 false negatives occurred within this single-criterion subgroup (Supplementary Table 2). The higher prevalence of mild deficits likely explains why the ensemble’s sensitivity (79%) was lower than its specificity (100%). Training future models to predict a continuous standardized measure of language impairment will provide more detailed predictions beyond a binary label, and ensure those with milder language impairments are not missed. Finally, expanding cohort scale and culture diversity will generalize these models across broader populations. Although this study represents the largest acute stroke cohort to date (N= 86; 63 with language impairment), prior work was smaller and restricted to stroke vs. control comparisons (e.g., Boucher et al., n =40 with LI; Bunker et al, n = 49 with LI)^16,17^. Evaluating larger, multi-center cohorts will further enhance classifier stability while testing narrative prompts beyond Western folklore (such as Cinderella) will ensure robust cross-cultural validity. Resolving these practical developments will bring automated connected speech analysis closer to routine acute care, reducing clinician workload and facilitating timely referrals to speech therapy.

In conclusion, this study developed a brief, 1-2-minute, largely automated screener of real-world speech that accurately identifies acute post-stroke language impairment from brief story retellings by combining automatic speech recognition, natural language processing (NLP)-derived linguistic features, large language model (LLM)-derived embeddings, and machine learning (ML)-based classification algorithms. An ensemble combining embeddings across diverse LLM architectures achieved 90% balanced accuracy, demonstrating that distinct language models capture complementary aspects of impaired discourse. Adding the linguistic classifier to the ensemble modestly increased sensitivity at the expense of specificity, a trade-off that may be appropriate when screening is used to prompt further language assessment and potential rehabilitation referral. By enabling individual-level differentiation within an acute post-stroke cohort, this work provides a foundation for prospective, externally validated discourse screening during acute stroke care.

## Materials and Methods

### Participants

#### Acute stroke participants

As part of a larger prospective project involving multiple comprehensive stroke centers in the Texas Medical Center in Houston, Texas, we consecutively recruited monolingual English speakers with acute left hemisphere stroke independent of clinical diagnosis of language impairment from April 27, 2011, to January 7, 2020. Written informed consent was obtained from either the patient or a legally authorized representative, as approved by the Institutional Review Boards at Baylor College of Medicine, Rice University, the University of Texas Health Sciences Center, and Houston Methodist Hospital.

A total of 101 participants completed the story retelling task (see Supplementary Figure 6 for inclusion/exclusion criteria and participant group assignment). We first excluded 6 participants with pre-existing health conditions affecting cognition (tumor (n=3); neurodegenerative disease (n=1); substance abuse disorder (n=2)). To establish ground-truth language status in the remaining cohort (N = 95), we implemented a composite reference standard using clinical data acquired independently from the story retelling task. Because formal, multi-subtest standardized language batteries, such as the Western Aphasia Battery^32^ (WAB), are rarely feasible during acute stroke care due to clinical restraints, we classified participants as having language impairment (LI) if they met at least one of three, independent criteria combining clinical and objective evidence: (i) a non-zero score on Item 9 (best language score) of the NIH stroke scale^5^, where scores of 0, 1, 2 and 3 indicate no, mild-to-moderate, severe and global language impairment respectively; (ii) an acute language impairment diagnosis documented by a speech-language pathologist (SLP) in electronic medical records (EMR); and (iii) a picture naming error > 10%^33^ derived from patient responses to 69 pictures of objects in a separate confrontation naming battery^12^. Participants viewed each picture on a computer screen and named the object with a single word within 10 seconds; confrontation naming accuracy strongly predicts aphasia severity^33^. Different from Schnur and Lei^12^, we categorized naming responses containing articulatory problems as errors.

We excluded 9 participants because language-impairment status could not be assigned with sufficient confidence. This group included 7 participants with intact naming (error rate ≤ 10%) who lacked both NIHSS Item 9 scores^5^ and EMR diagnosis, precluding definitive classification regarding the presence or absence of language impairment (e.g., dynamic aphasia, an impairment of discourse but intact naming)^34^ and 2 participants for whom data across all three criteria were unavailable.

Following these exclusions, the final sample comprised 86 acute stroke participants (63 with LI, 23 with no LI; 35 females, 11 left-handed, 2 ambidextrous; average age of 61 years, range = 25-85 years; average education of 14 years, range = 6-33 years; average naming error of 15%, range = 0-93%; average Item 9 score of 0.26, range 0-2). Among the 63 participants classified with LI, five patients met all three diagnostic criteria, 18 met two criteria, and 40 met a single criterion. Diagnostic distributions and missing data parameters across these criteria are summarized in Supplementary Table 1.

Evaluations were completed entirely within the acute window. Story retelling and confrontation naming tasks were administered at an average of 4 days post-stroke (range 1-17 days). SLP evaluations documented in the EMR occurred at an average of 3 days post-stroke (range 0-16 days). NIHSS Item 9 screening occurred at an average of 2 days post-stroke (range 0-10 days) and within close temporal proximity to behavioral testing (average = 0.2 days, range 0-1 days).

#### Control participants

Twenty neurologically healthy control participants (10 female; 1 left-handed) were matched to the patient sample for age (mean = 56 years, range 32-77 years; t = 1.6, p = 0.105) and education (mean = 14 years, range 12-20; t = 0.03, p = 0.97). Control participants reported no history of neurological disorders, no significant visual or auditory impairments, and were screened against cognitive impairment using the Montreal Cognitive Assessment (MoCA)^36^ with an alternative cut-off score of 23 to reduce the demographic bias^37^. Story retellings from controls were used to derive global coherence baseline values for the patient cohort (see Supplementary Table 3).

### Story retelling

Participants viewed a picture book of the Cinderella story^38^ for as long as they wanted, with the words concealed. Participants were then instructed to close the book and retell the story of Cinderella. Experimenters encouraged participants to produce full sentences and continue speaking if they produced long hesitations mid-narrative.

### Procedure

Participants were evaluated either in the hospital or at their homes if they were discharged before testing was completed. We administered and audio-recorded picture naming followed by story retellings^8,10,12^.

### Preprocessing and transcription

We manually removed non-participant speech and silence at the start and end of audio recordings. For automated story retelling transcription, we applied the ASR tool Whisper large-v3^39^ to the recordings. We chose Whisper because Whisper-generated transcriptions of connected speech samples accurately classify language impairment at the chronic stage of stroke^15^.

### Deriving discrete linguistic features

From the transcribed story retellings, we derived a total of 14 discrete linguistic features previously demonstrated to be sensitive to discourse deficits in acute or chronic stroke-induced language impairment^10,15,40-47^. Features included those derived at the level of discourse (n=3), utterance (n=2), and word (n=9). See Supplementary Table 3. We included word-level derived features when values were available for >80% of words across all patients’ narratives. Features were derived with code adapted from previous studies^28,48,49^.

### Deriving embeddings

We derived embeddings using GloVe^22^ and three LLMs (BERT^23^, Mistral^24^, and OpenAI^25^), which have successfully differentiated picture descriptions produced by participants with Alzheimer’s disease from neurologically healthy controls^18,19^. For GloVe, we derived 300-dimensional pre-trained vectors from the largest GloVe model trained for each word in a given narrative. We averaged each word’s vector to yield a 300-dimensional GloVe-based representation for a given narrative. For BERT, we applied the bert-large-cased model from the HuggingFace library, using the 1024-dimensional vector representation of the CLS token, a special token that served as the vector representation of the entire narrative^50^. For Mistral, we derived a 4096-dimensional vector representation for each narrative using the Linq-AI-Research/Linq-Embed-Mistral from the Hugging Face library. For OpenAI, we derived a 3072-dimensional vector representation for each narrative using the text-embedding-3-large model.

### Classifier design

For an overview of the methodological approach, see Figure 3.

**Figure 3.**
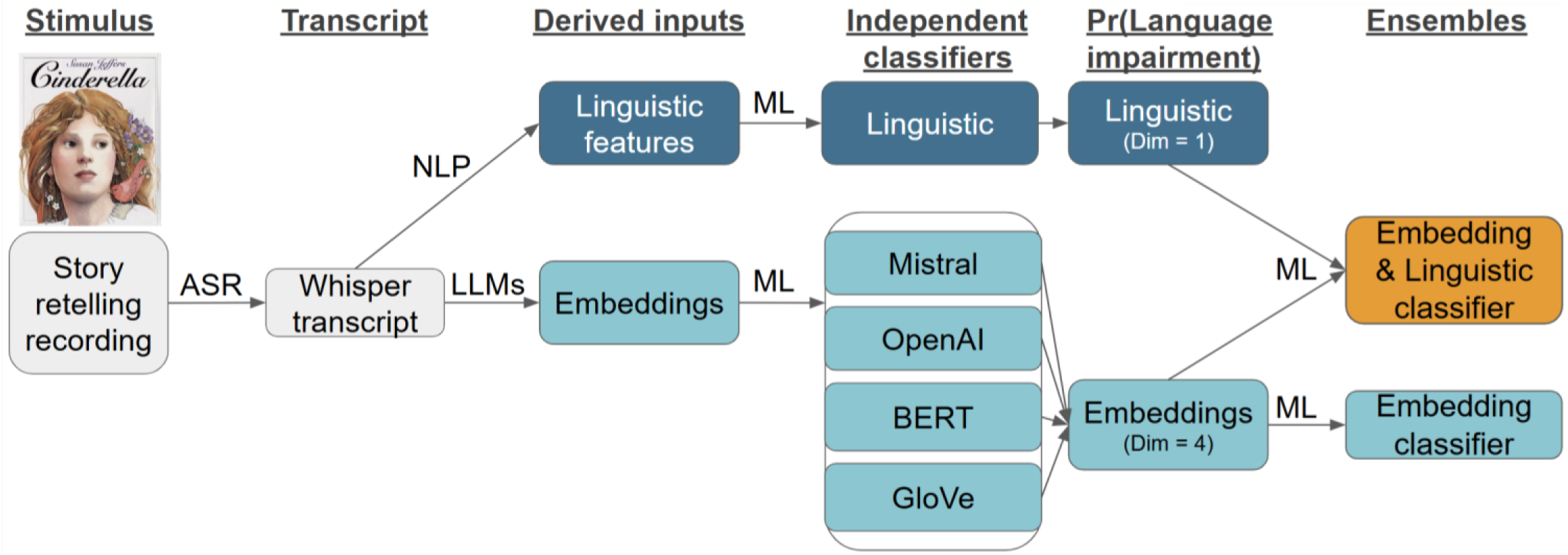
Summary of methodological approach. First, Cinderella story retelling^38^ recordings were transcribed by the automatic speech recognition (ASR) tool Whisper^39^. Then, we used natural language processing (NLP), large language models (LLMs)^23-25^ (Mistral, OpenAI, BERT) and GloVe^22^ to derive discrete linguistic features and embeddings. Machine learning (ML) was used to create a classifier for discrete linguistic features, a classifier for GloVe embeddings, and classifiers for each LLM’s embedding. From each embedding-based classifier, we derived language-impairment prediction probabilities, denoted as Pr(Language Impairment), to construct an embedding-based ensemble classifier^20,51^. From the linguistic and embedding-based classifiers, we derived language-impairment prediction probabilities to construct an embedding-and-linguistic-based ensemble classifier. ASR = Automatic speech recognition, NLP = Natural language processing, LLMs = Large language models, ML = machine learning, Dim = Dimension, Pr = Probability. Key: Grey = Patient data, Dark blue = Linguistic, Light blue = Embeddings, Orange = Embedding & Linguistic.

### Classification algorithms

Due to the lack of a current consensus on the best classification algorithm given discrete linguistic features or embeddings as input, following previous studies, we reported the performance of multiple classification algorithms^13,15,18,52-55,^. We included classification algorithms that demonstrated above-chance classification performance using discrete linguistic features or embeddings as input in solving similar classification tasks in clinical populations with chronic language impairments: shallow neural network^53^, decision tree^13,15,52,53^, support vector machine^13-15,18,52-55^, logistic regression^18,19,54^, random forest ^13,15,18,21,52,55^, and gradient boosting^15,56^. We evaluated these classification algorithms using the ScikitLearn^57^ Python package. As support vector machines and shallow neural networks assume that each feature is on the same scale, we z-scored our data before evaluating these two classification algorithms. After z-scoring, principal component analysis was applied, where the number of principal components was treated as a hyperparameter to be tuned. For the shallow neural network classification algorithm, we used only one hidden layer to avoid overfitting due to the size of our sample.

#### Hyperparameter tuning and evaluation

We adopted nested leave-one-out cross-validation (LOOCV) as it provides an unbiased estimate of true generalization performance based on the size of our sample (sample sizes of < 100)^58^. We used five inner folds for hyperparameter tuning. We used *n* outer folds (*n* being # of samples, i.e., leave-one-out) to ensure that the reported results were applicable across participants. Classifier construction, hyperparameter tuning, and LOOCV were automated using the Python package ScikitLearn^57^. For each classifier, we employed Receiver Operating Characteristic (ROC) analysis to determine the optimal classification threshold, following the approach outlined by Bedrick and colleagues^59^. To confirm that classification performance was not dependent on this threshold, we evaluated whether the classifier’s balanced accuracy was comparable (within 10%) to the classifier’s area-under-curve score, which represents the predictive power of a classifier without the need for a specific threshold^56^.

To evaluate the success of a classification algorithm, we first derived a confusion matrix indicating the number of true positives (TP), true negatives (TN), false positives (FP), and false negatives (FN). Confusion matrices were derived with ScikitLearn’s confusion_matrix function. From this confusion matrix, we calculated each classification algorithm’s balanced accuracy, defined as the average of specificity and sensitivity, where Sensitivity = TP / (TP + FN) (i.e., % of participants with language impairment correctly classified as having language impairment), and Specificity = TN / (TN + FP) (i.e., % of participants without language impairment correctly classified as not having language impairment). To compute the confidence interval of each evaluation metric for a given classifier, we used the bootstrap_ci method of the Python package confidenceinterval^60^, with number of resamples set to 10000.

To evaluate whether a classification algorithm performed above chance, we compared its classification performance against a naive classifier (the most frequent class baseline), which predicted every sample as the most common class in our sample (i.e., language impairment), implemented through ScikitLearn’s DummyClassifier model, with the strategy parameter set to “most_frequent.”

#### Selection of classification algorithm and ensembling

To select each feature set’s (discrete linguistic features, GloVe/BERT/Mistral/OpenAI embeddings) classification algorithm for use in the ensemble modeling step, we chose the ML-based classification algorithm with the highest balanced accuracy, assuming its AUC score was no more than 10% lower than its balanced accuracy score. For example, if the support vector machine classification algorithm achieved the highest balanced accuracy score given BERT embeddings, and its AUC score was no more than 10% lower than its balanced accuracy score, the final BERT embedding classifier will be a support vector machine. This followed previous language impairment classification studies that evaluated multiple classification algorithms and reported the performance of the most accurate^13,15,18,52-55^. Then, the language impairment prediction probabilities from each feature set’s final classifier were included as input to the classification algorithms to build two late-fusion^20^ ensemble models (Figure 3), following Reda et al.^51^. The first ensemble model consisted of the four embedding-based classifiers. The second ensemble model consisted of the linguistic classifier and the four embedding-based classifiers. Like the final classifier for each feature set, each ensemble model was defined as the ML-based classification algorithm that achieved the highest balanced accuracy score given the ensemble’s input, assuming its AUC score was no more than 10% lower than its balanced accuracy score.

#### Feature importance

For the linguistic classifier, we conducted feature-permutation analysis to identify the most important discrete linguistic features for classification success. For each feature, we calculated the average decrease in classification performance from 10000 random permutations of that feature. Feature permutation analysis was conducted with ScikitLearn’s *permutation_importance* method^20,57^. For the ensemble classifier, we conducted feature permutation analysis to compute the importance of each independent classifier. For each classifier’s language impairment prediction probability, we calculated the average decrease in the ensemble’s classification performance from 10000 random permutations of that feature.

## Acknowledgements

We would like to thank Miranda Brenneman, Cris Hamilton, Chia-Ming Lei, and Danielle Rossi for their help in data collection. We would like to thank Emilia Cichocki, Erica Johns, Bowie Lin, Hao Yan, and Rachel Zahn for their help in transcription and analysis. We would like to thank the ICU teams at Memorial Hermann Hospital, Houston Methodist Hospital, and Baylor St. Luke’s Hospital. We would like to thank Danielle Brown, speech-language pathologist, for sharing her clinical expertise. We would like to thank our patient coordinator, Andrea Suazo, for her feedback. We would like to thank Hanjie Chen for her suggestion of evaluating multiple LLMs. We would like to thank Eric Wuesthoff for their support with science communication. Portions of this work were presented at the 2025 Cognitive Neuroscience Society. Lastly, we would like to thank our participants and their caregivers for their time and effort, which made this work possible.

## Author contributions

Lokesha Pugalenthi: Conceptualization, Data curation, Formal analysis, Methodology, Writing-Original Draft, Writing-Review & Editing Tatiana Schnur: Conceptualization, Investigation, Resources, Data Curation, Writing-Review & Editing, Funding acquisition, Project administration, Supervision

## Data availability

The data supporting the findings of this study are subject to HIPAA regulations. However, non-identifying data will be shared upon reasonable request based on a formal data-sharing agreement. Please contact the corresponding author, Tatiana Schnur, for data use inquiries at. Analytical code and intermediary outputs are available at https://github.com/lpugalenthi/pugalenthiSchnurPaper2026ms/.

## References

1. Cruice, M., Worrall, L., Hickson, L., & Murison, R. (2003). Finding a focus for quality of life with aphasia: Social and emotional health, and psychological well-being. Aphasiology, 17(4), 333–353.

2. Wallace, S. J., Worrall, L., Rose, T., Le Dorze, G., Cruice, M., Isaksen, J., … & Gauvreau, C. A. (2017). Which outcomes are most important to people with aphasia and their families? An international nominal group technique study framed within the ICF. Disability and rehabilitation, 39(14), 1364–1379.

3. Hallin, A. E., & Partanen, P. (2024). Factors affecting speech-language pathologists’ language assessment procedures and tools – challenges and future directions in Sweden. Logopedics Phoniatrics Vocology, 49(3), 104–113. 10.1080/14015439.2022.2158218

4. Stark, B. C., Dutta, M., Murray, L. L., Fromm, D., Bryant, L., Harmon, T. G., Ramage, A. E., & Roberts, A. C. (2021). Spoken Discourse Assessment and Analysis in Aphasia: An International Survey of Current Practices. Journal of Speech, Language, and Hearing Research, 64(11), 4366–4389. 10.1044/2021_JSLHR-20-00708

5. Kwah, L. K., & Diong, J. (2014). National institutes of health stroke scale (NIHSS). Journal of Physiotherapy, 60(1), 61.

6. Fromm, D., Forbes, M., Holland, A., Dalton, S. G., Richardson, J., & MacWhinney, B. (2017). Discourse Characteristics in Aphasia Beyond the Western Aphasia Battery Cutoff. American Journal of Speech-Language Pathology, 26(3), 762–768. 10.1044/2016_AJSLP-16-0071

7. Fromm, D., Katta, S., Paccione, M., Hecht, S., Greenhouse, J., MacWhinney, B., & Schnur, T. T. (2021). A Comparison of Manual Versus Automated Quantitative Production Analysis of Connected Speech. Journal of Speech, Language, and Hearing Research, 64(4), 1271–1282. 10.1044/2020_JSLHR-20-00561

8. Schnur, T. T., & Wang, S. (2024). Differences in Connected Speech Outcomes Across Elicitation Methods. Aphasiology, 38(5), 816–837. 10.1080/02687038.2023.2239509

9. Clarke, N., Barrick, T. R., & Garrard, P. (2021). A Comparison of Connected Speech Tasks for Detecting Early Alzheimer’s Disease and Mild Cognitive Impairment Using Natural Language Processing and Machine Learning. Frontiers in Computer Science, 3. 10.3389/fcomp.2021.634360

10. Ding, J., Martin, R. C., Hamilton, A. C., & Schnur, T. T. (2020). Dissociation between frontal and temporal-parietal contributions to connected speech in acute stroke. Brain, 143(3), 862–876.

11. Martin, R. C., & Schnur, T. T. (2019). Independent contributions of semantic and phonological working memory to spontaneous speech in acute stroke. Cortex, 112, 58–68.

12. Schnur, T. T., & Lei, C.-M. (2022). Assessing naming errors using an automated machine learning approach. Neuropsychology, 36(8), 709–718. 10.1037/neu0000860

13. Chatzoudis, G., Plitsis, M., Stamouli, S., Dimou, A.-L., Katsamanis, A., & Katsouros, V. (2022). Zero-Shot Cross-lingual Aphasia Detection using Automatic Speech Recognition (arXiv:2204.00448). arXiv. 10.48550/arXiv.2204.00448

14. Zusag, M., Wagner, L., & Bloder, T. (2023). Careful Whisper-leveraging advances in automatic speech recognition for robust and interpretable aphasia subtype classification. Proc. Interspeech 2023, 3013– 3017. https://www.isca-archive.org/interspeech_2023/zusag23_interspeech.html

15. Cong, Y., LaCroix, A. N., & Lee, J. (2024). Clinical efficacy of pre-trained large language models through the lens of aphasia. Scientific Reports, 14(1), 15573.

16. Boucher, J., Marcotte, K., Brisebois, A., Courson, M., Houzé, B., Desautels, A., Léonard, C., Rochon, E., & Brambati, S. M. (2022). Word-finding in confrontation naming and picture descriptions produced by individuals with early post-stroke aphasia. The Clinical Neuropsychologist, 36(6), 1422–1437. 10.1080/13854046.2020.1817563

17. Bunker, L. D., Berube, S. K., Neal, V., Kelly, L., Kelly, C., Meier, E. L., & Hillis, A. E. (2025). Discourse Measures From the Modern Cookie Theft Picture Description Are Sensitive to Mild Communication Deficits Not Captured by the Western Aphasia Battery–Revised Aphasia Quotient. American Journal of Speech-Language Pathology, 34(3), 1100–1120. 10.1044/2024_AJSLP-24-00322

18. Agbavor, F., & Liang, H. (2022). Predicting dementia from spontaneous speech using large language models. PLOS Digital Health, 1(12), e0000168. 10.1371/journal.pdig.0000168

19. Llaca-Sánchez, B. A., García-Noguez, L. R., Aceves-Fernández, M. A., Takacs, A., & Tovar-Arriaga, S. (2025). Exploring LLM Embedding Potential for Dementia Detection Using Audio Transcripts. Eng, 6(7). 10.3390/eng6070163

20. Huang, S.-C., Pareek, A., Seyyedi, S., Banerjee, I., & Lungren, M. P. (2020). Fusion of medical imaging and electronic health records using deep learning: A systematic review and implementation guidelines. NPJ Digital Medicine, 3(1), 136.

21. Breiman, L. (2001). Random Forests. Machine Learning, 45(1), 5–32. 10.1023/A:1010933404324

22. Pennington, J., Socher, R., & Manning, C. (2014). GloVe: Global Vectors for Word Representation. In A. Moschitti, B. Pang, & W. Daelemans (Eds.), Proceedings of the 2014 Conference on Empirical Methods in Natural Language Processing (EMNLP) (pp. 1532–1543). Association for Computational Linguistics. 10.3115/v1/D14-1162

23. Devlin, J., Chang, M.-W., Lee, K., & Toutanova, K. (2019). BERT: Pre-training of Deep Bidirectional Transformers for Language Understanding. In J. Burstein, C. Doran, & T. Solorio (Eds.), Proceedings of the 2019 Conference of the North American Chapter of the Association for Computational Linguistics: Human Language Technologies, Volume 1 (Long and Short Papers) (pp. 4171–4186). Association for Computational Linguistics. 10.18653/v1/N19-1423

24. Choi, C., Kim, J., Lee, S., Kwon, J., Gu, S., Kim, Y., … & Sohn, J. Y. (2024). Linq-embed-mistral technical report. arXiv preprint arXiv:2412.03223.

25. Text-embedding-3-large Model | OpenAI API. (2024). Retrieved March 15, 2026, from https://developers.openai.com/api/docs/models/text-embedding-3-large

26. Domor, M. I., & Swart, T. G. (2025). Ensemble Large Language Models: A Survey. Information, 16(8), 688–711. 10.3390/info16080688

27. Plaquet, A., & Bredin, H. (2023). Powerset multi-class cross entropy loss for neural speaker diarization. 3222–3226. 10.21437/Interspeech.2023-205

28. Hilsabeck, R. C., Keller, J. N., Henry, M. L., Li, J. J., Pugalenthi, L., Toprac, P., … & Rathouz, P. J. (2025). Development and classification accuracy of an automated cognitive screening tool combining working memory and connected speech tasks for early detection of cognitive impairment in primary care. Alzheimer’s & Dementia: Translational Research & Clinical Interventions, 11(3), e70145.

29. Assis, A., Dantas, J., & Andrade, E. (2024). The performance-interpretability trade-off: A comparative study of machine learning models. Journal of Reliable Intelligent Environments, 11(1), 1. 10.1007/s40860-024-00240-0

30. Lundberg, S. M., & Lee, S.-I. (2017). A Unified Approach to Interpreting Model Predictions. Advances in Neural Information Processing Systems, 30. https://proceedings.neurips.cc/paper/2017/hash/8a20a8621978632d76c43dfd28b67767-Abstract.html

31. Merhbene, G., Lecron, F., Fortemps, P., Dickerson, B. C., Kurpicz-Briki, M., & Rezaii, N. (2026). Detecting Primary Progressive Aphasia (PPA) from Text: A Benchmarking Study. In V. Demberg, K. Inui, & L. Marquez (Eds.), Findings of the Association for Computational Linguistics: EACL 2026 (pp. 355–374). Association for Computational Linguistics. 10.18653/v1/2026.findings-eacl.19

32. Kertesz, A. (2007). Western aphasia battery–revised. https://psycnet.apa.org/doiLanding?doi=10.1037/t15168-000

33. Hillis, A. E., Wityk, R. J., Barker, P. B., Beauchamp, N. J., Gailloud, P., Murphy, K., Cooper, O., & Metter, E. J. (2002). Subcortical aphasia and neglect in acute stroke: The role of cortical hypoperfusion. Brain, 125(5), 1094–1104. 10.1093/brain/awf113

34. Walker, G. M., Fridriksson, J., Hillis, A. E., Den Ouden, D. B., Bonilha, L., & Hickok, G. (2022). The Severity-Calibrated Aphasia Naming Test. American Journal of Speech-Language Pathology, 31(6), 2722–2740. 10.1044/2022_AJSLP-22-00071

35. Robinson, G., Blair, J., & Cipolotti, L. (1998). Dynamic aphasia: An inability to select between competing verbal responses? Brain, 121(1), 77–89. 10.1093/brain/121.1.77

36. Nasreddine, Z. S., Phillips, N. A., Bédirian, V., Charbonneau, S., Whitehead, V., Collin, I., Cummings, J. L., & Chertkow, H. (2005). The Montreal Cognitive Assessment, MoCA: A brief screening tool for mild cognitive impairment. Journal of the American Geriatrics Society, 53(4), 695–699. 10.1111/j.1532-5415.2005.53221.x

37. Carson, N., Leach, L., & Murphy, K. J. (2018). A re-examination of Montreal Cognitive Assessment (MOCA) cutoff scores. International Journal of Geriatric Psychiatry, 33(2), 379–388. 10.1002/gps.4756

38. Jeffers, S., Perrault, C., & Ehrlich, A. (2004). Cinderella. Dutton Children’s Books.

39. Radford, A., Kim, J. W., Xu, T., Brockman, G., Mcleavey, C., & Sutskever, I. (2023). Robust Speech Recognition via Large-Scale Weak Supervision. Proceedings of the 40th International Conference on Machine Learning, 28492–28518. https://proceedings.mlr.press/v202/radford23a.html

40. Alyahya, R. S., Lambon Ralph, M. A., Halai, A., & Hoffman, P. (2022). The cognitive and neural underpinnings of discourse coherence in post-stroke aphasia. Brain Communications, 4(3), fcac147.

41. Brysbaert, M., & Ellis, A. W. (2016). Aphasia and age of acquisition: Are early-learned words more resilient? Aphasiology, 30(11), 1240–1263. 10.1080/02687038.2015.1106439

42. Cunningham, K. T., & Haley, K. L. (2020). Measuring Lexical Diversity for Discourse Analysis in Aphasia: Moving-Average Type–Token Ratio and Word Information Measure. Journal of Speech, Language, and Hearing Research, 63(3), 710–721. 10.1044/2019_JSLHR-19-00226

43. Saffran, E. M., Berndt, R. S., & Schwartz, M. F. (1989). The quantitative analysis of agrammatic production: Procedure and data. Brain and Language, 37(3), 440–479.

44. Stark, B. C., Dalton, S. G., & Lanzi, A. M. (2025). Access to context-specific lexical-semantic information during discourse tasks differentiates speakers with latent aphasia, mild cognitive impairment, and cognitively healthy adults. Frontiers in Human Neuroscience, 18. 10.3389/fnhum.2024.1500735

45. Hoffman, P., Cogdell-Brooke, L., & Thompson, H. E. (2020). Going off the rails: Impaired coherence in the speech of patients with semantic control deficits. Neuropsychologia, 146, 107516. 10.1016/j.neuropsychologia.2020.107516

46. Litovsky, C. P., Finley, A. M., Zuckerman, B., Sayers, M., Schoenhard, J. A., Kenett, Y. N., & Reilly, J. (2022). Semantic flow and its relation to controlled semantic retrieval deficits in the narrative production of people with aphasia. Neuropsychologia, 170, 108235.

47. Mancano, M., & Papagno, C. (2023). Concrete and Abstract Concepts in Primary Progressive Aphasia and Alzheimer’s Disease: A Scoping Review. Brain Sciences, 13(5), 765. 10.3390/brainsci13050765

48. Misra, K. (2022). minicons: Enabling flexible behavioral and representational analyses of transformer language models. arXiv preprint arXiv:2203.13112.

49. Hoffman, P., Loginova, E., & Russell, A. (2018). Poor coherence in older people’s speech is explained by impaired semantic and executive processes. eLife, 7, e38907. 10.7554/eLife.38907

50. Rogers, A., Kovaleva, O., & Rumshisky, A. (2020). A Primer in BERTology: What We Know About How BERT Works. Transactions of the Association for Computational Linguistics, 8, 842–866. 10.1162/tacl_a_00349

51. Reda, I., Khalil, A., Elmogy, M., Abou El-Fetouh, A., Shalaby, A., Abou El-Ghar, M., Elmaghraby, A., Ghazal, M., & El-Baz, A. (2018). Deep Learning Role in Early Diagnosis of Prostate Cancer. Technology in Cancer Research & Treatment, 17, 1533034618775530. 10.1177/1533034618775530

52. Themistocleous, C., Ficek, B., Webster, K., Den Ouden, D.-B., Hillis, A. E., & Tsapkini, K. (2021). Automatic Subtyping of Individuals with Primary Progressive Aphasia. Journal of Alzheimer’s Disease, 79(3), 1185–1194. 10.3233/JAD-201101

53. Matias-Guiu, J. A., Suárez-Coalla, P., Yus, M., Pytel, V., Hernández-Lorenzo, L., Delgado-Alonso, C., Delgado-Álvarez, A., Gómez-Ruiz, N., Polidura, C., Cabrera-Martín, M. N., Matías-Guiu, J., & Cuetos, F. (2022). Identification of the main components of spontaneous speech in primary progressive aphasia and their neural underpinnings using multimodal MRI and FDG-PET imaging. Cortex, 146, 141–160. 10.1016/j.cortex.2021.10.010

54. Fraser, K. C., Meltzer, J. A., Graham, N. L., Leonard, C., Hirst, G., Black, S. E., & Rochon, E. (2014). Automated classification of primary progressive aphasia subtypes from narrative speech transcripts. *Cortex, Language*, Computers and Cognitive Neuroscience, 55, 43–60. 10.1016/j.cortex.2012.12.006

55. Balagopalan, A., Novikova, J., Mcdermott, M. B., Nestor, B., Naumann, T., & Ghassemi, M. (2020). Cross-language aphasia detection using optimal transport domain adaptation. Machine Learning for Health Workshop, 202–219. http://proceedings.mlr.press/v116/balagopalan20a.html

56. Lukic, S., Fan, Z., García, A. M., Welch, A. E., Ratnasiri, B. M., Wilson, S. M., Henry, M. L., Vonk, J., Deleon, J., & Miller, B. L. (2024). Discriminating nonfluent/agrammatic and logopenic PPA variants with automatically extracted morphosyntactic measures from connected speech. Cortex, 173, 34–48.

57. Pedregosa, F., Varoquaux, G., Gramfort, A., Michel, V., Thirion, B., Grisel, O., … & Cournapeau, D. (2011). Scikit-learn: Machine learning in Python. machine learning in python, 6.

58. Wong, T.-T. (2015). Performance evaluation of classification algorithms by k-fold and leave-one-out cross validation. Pattern Recognition, 48(9), 2839–2846. 10.1016/j.patcog.2015.03.009

59. Bedrick, S. (2024, August 23). Computational analysis of aphasic narrative speech: Are we there yet? http://cstar.sc.edu/lecture-series/

60. Python library for confidence intervals (2023). Retrieved July 15, 2026, from https://github.com/jacobgil/confidenceinterval

61. Jiang, A. Q., Sablayrolles, A., Mensch, A., Bamford, C., Chaplot, D. S., Casas, D. de las, Bressand, F., Lengyel, G., Lample, G., Saulnier, L., Lavaud, L. R., Lachaux, M.-A., Stock, P., Scao, T. L., Lavril, T., Wang, T., Lacroix, T., & Sayed, W. E. (2023). Mistral 7B (arXiv:2310.06825). arXiv. 10.48550/arXiv.2310.06825

62. Hoffman, P., Lambon Ralph, M. A., & Rogers, T. T. (2013). Semantic diversity: A measure of semantic ambiguity based on variability in the contextual usage of words. Behavior Research Methods, 45(3), 718–730. 10.3758/s13428-012-0278-x

63. Kuperman, V., Stadthagen-Gonzalez, H., & Brysbaert, M. (2012). Age-of-acquisition ratings for 30,000 English words. Behavior Research Methods, 44(4), 978–990. 10.3758/s13428-012-0210-4

64. Brysbaert, M., New, B. Moving beyond Kučera and Francis: A critical evaluation of current word frequency norms and the introduction of a new and improved word frequency measure for American English. Behavior Research Methods 41, 977–990 (2009). 10.3758/BRM.41.4.977

65. Brysbaert, M., Warriner, A. B., & Kuperman, V. (2014). Concreteness ratings for 40 thousand generally known English word lemmas. Behavior Research Methods, 46(3), 904–911. 10.3758/s13428-013-0403-5

