## Supplementary Materials for "Automated language impairment screening in acute stroke using connected speech"

### 5. Supplementary Materials

#### Supplementary Figures and Figure Legends

**Supplementary Figure 1. Classification performance and confidence intervals for the linguistic, embedding-only ensemble, and embedding-and-linguistic ensemble classifiers.** Comparison of evaluation metrics (balanced accuracy, sensitivity, specificity, accuracy, and area under the receiver operating characteristic curve (AUC)). Error bars represent 95% confidence intervals computed using 10,000 bootstrap resamples via the Python confidenceinterval package[60]. AUC values multiplied by 100 for display consistency. Sensitivity is the proportion of participants with language impairment correctly identified; specificity is the proportion of participants without language impairment correctly identified. Balanced accuracy is the average of sensitivity and specificity. Accuracy is the proportion of participants correctly identified. Chance-level performance for balanced accuracy and AUC is 50%.

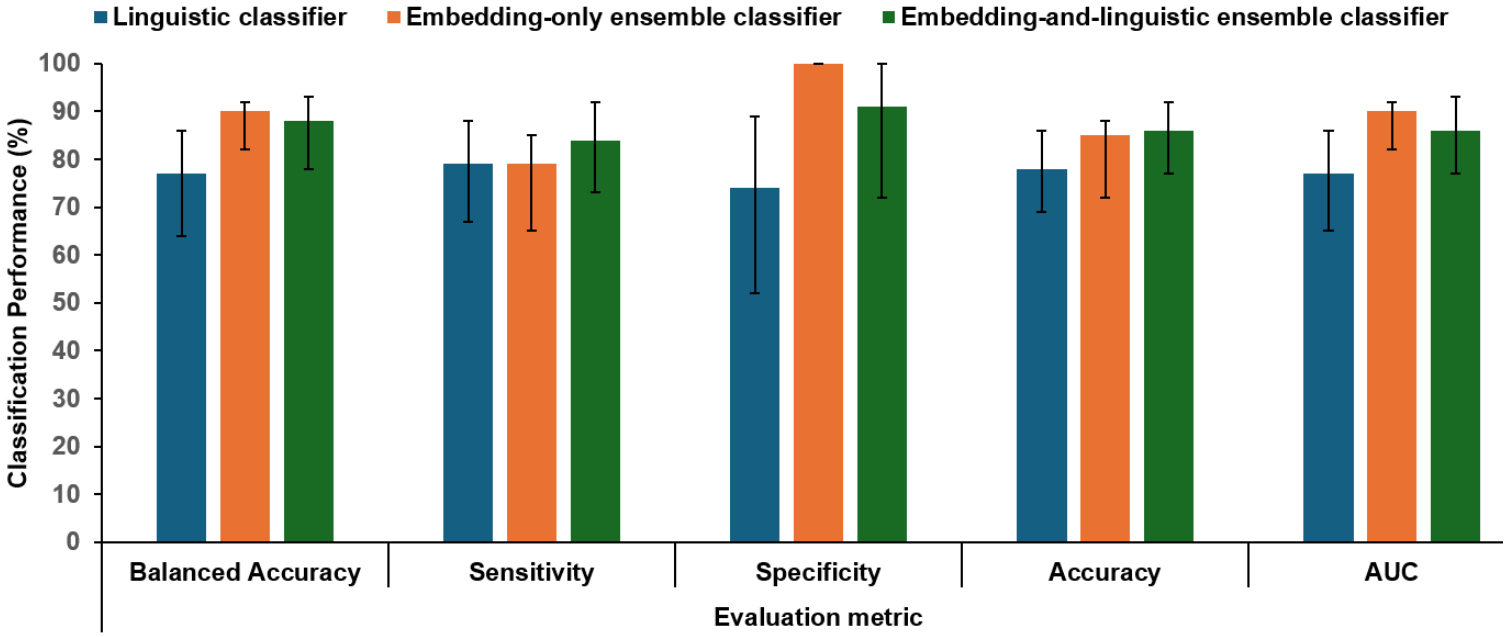

**Supplementary Figure 2. Feature importance for the linguistic classifier.** Permutation feature importance for the logistic regression model trained on the linguistic feature set. Bar lengths represent mean % drop in classification performance when individual feature values were randomly permuted. Only features causing a ≥2% decrease in classification performance are shown. Rounded values displayed to the right of each bar.

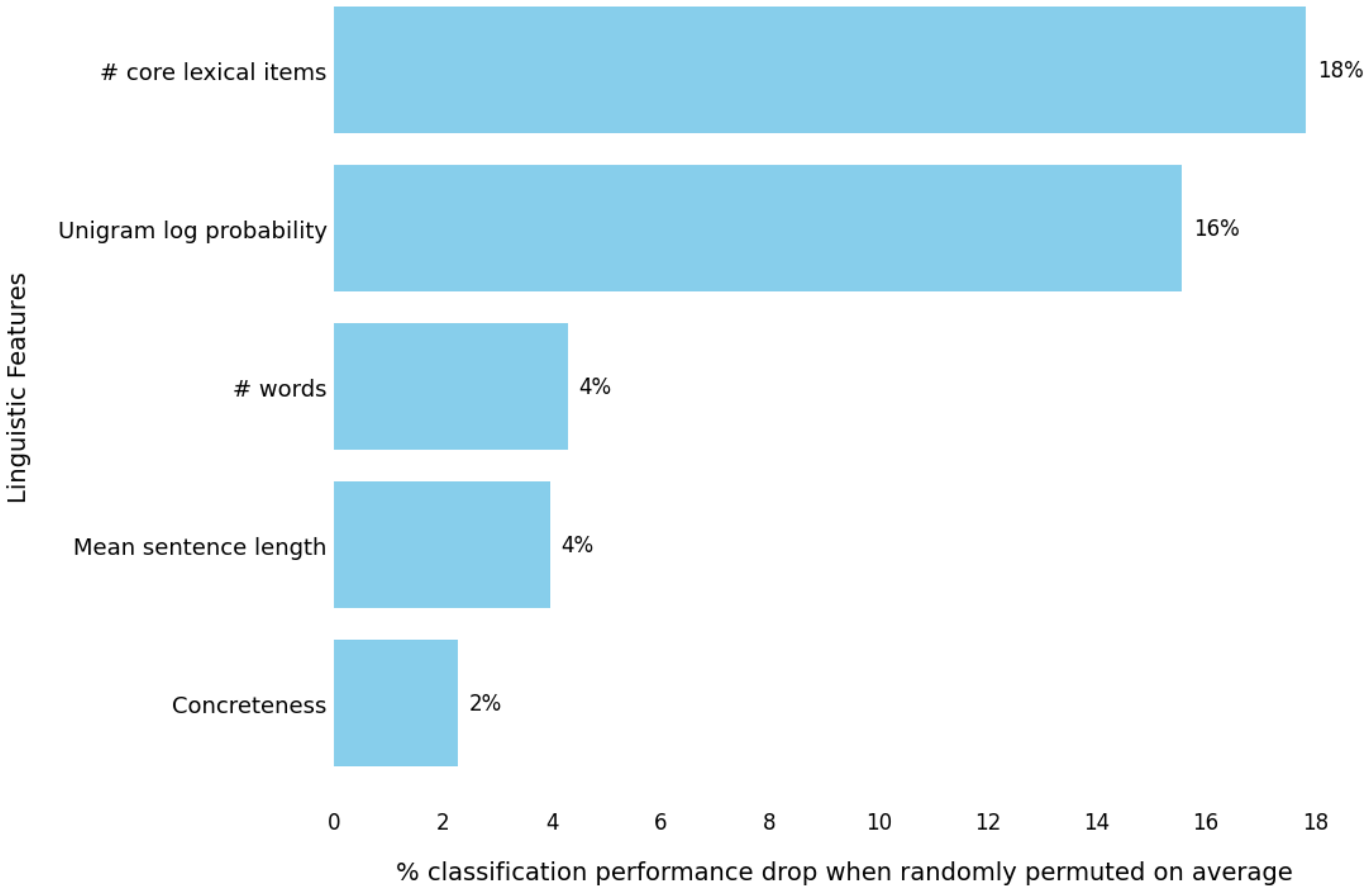

**Supplementary Figure 3. Distributions of linguistic feature values in stroke participants with and without language impairment.** Violin plots (a-n) illustrate the distributions of feature values for the linguistic classifier in participants with language impairment (n = 63, red) and without language impairment (n = 23, yellow). Panels are ordered by feature importance. Black dots indicate individual participant values; dashed lines indicate quartiles.

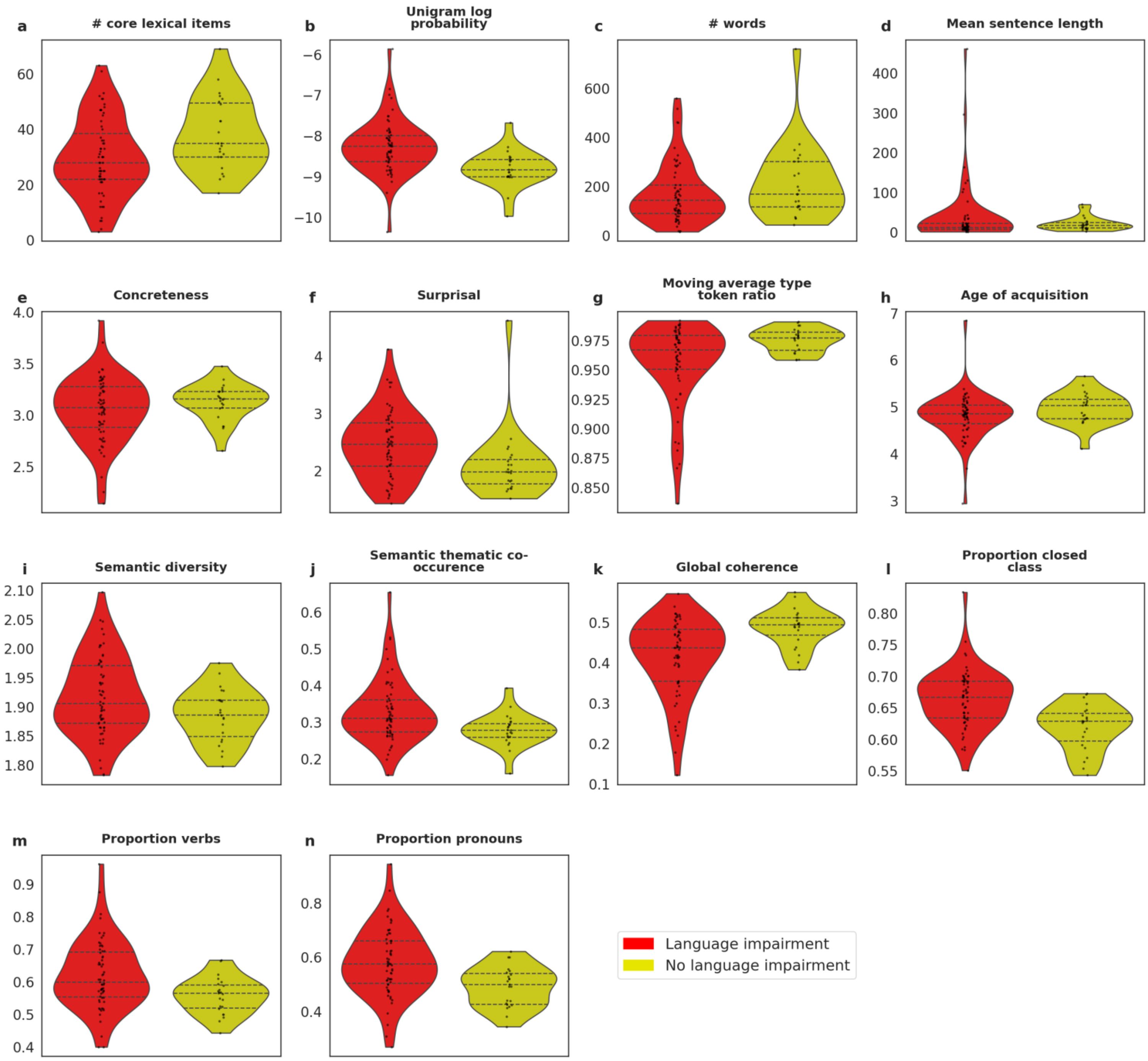

**Supplementary Figure 4. Classification performance across candidate machine learning algorithms for each classifier.** Evaluation metrics across candidate classification algorithms trained, ordered by highest balanced accuracy (ties broken by Area Under the Receiver Operating Characteristic Curve (AUC), for the linguistic classifier (**a**), embedding-only ensemble classifier (**b**), embedding-and-linguistic ensemble classifier (**c**). Values are shown as percentages; AUC values multiplied by 100 for display consistency. Sensitivity represents the proportion of participants with language impairment correctly identified; specificity represents the proportion of participants without language impairment correctly identified. Balanced accuracy is the average of sensitivity and specificity. Accuracy is the proportion of participants correctly identified. Chance-level performance for balanced accuracy and AUC is 50%. *Decision Tree was not designated as the top-performing algorithm for the linguistic classifier because its AUC was >10% lower than its balanced accuracy score.

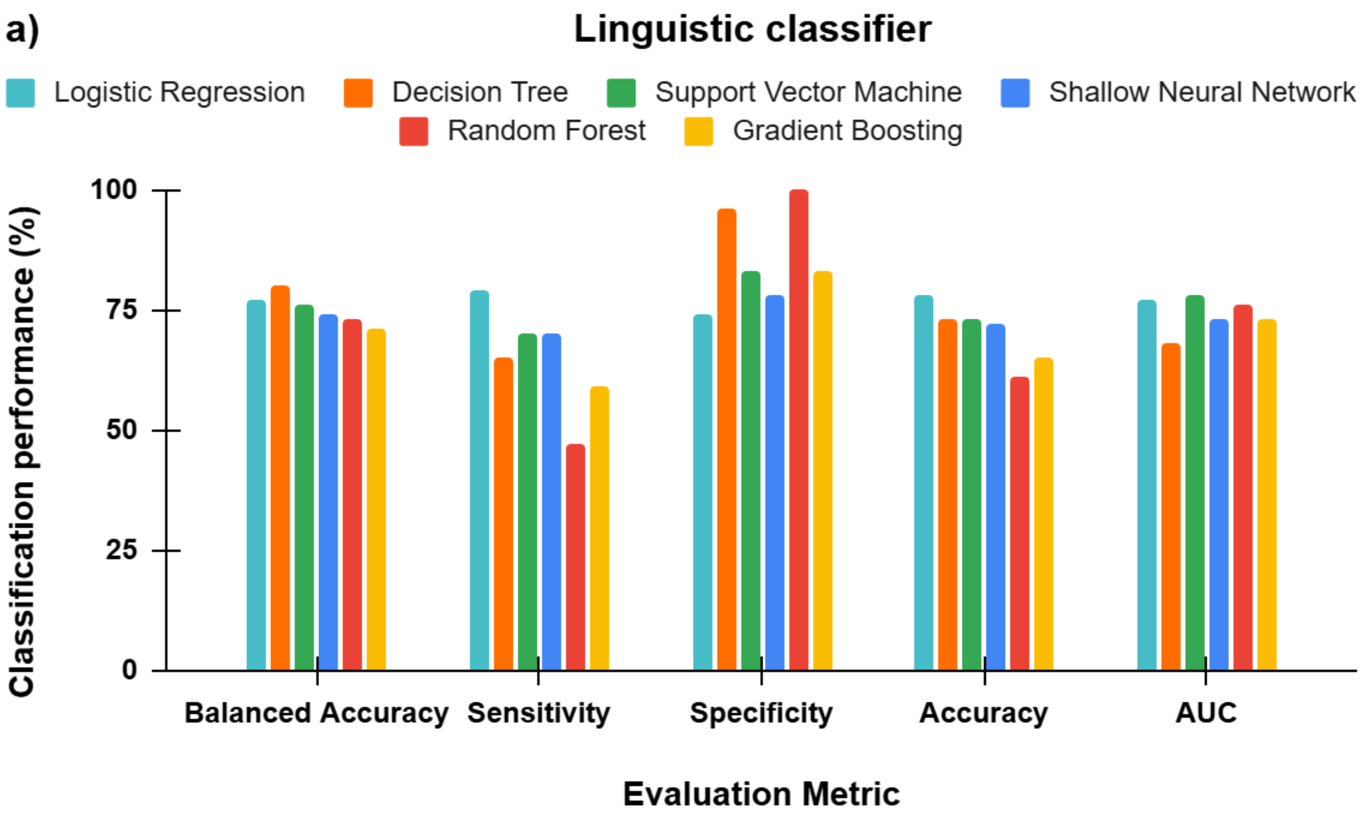

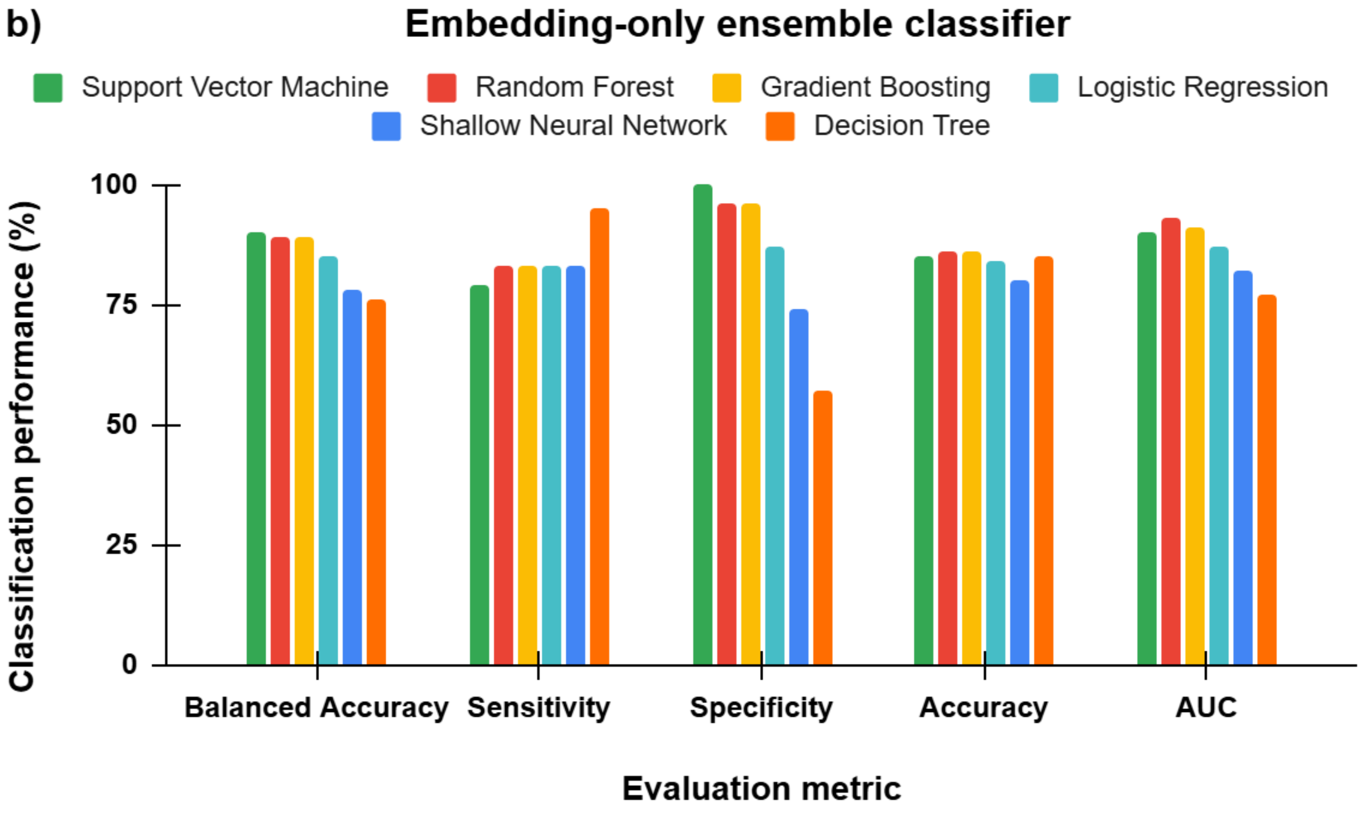
b)
Embedding-only ensemble classifier
Support Vector Machine
Random Forest
Gradient Boosting
Logistic Regression
Shallow Neural Network
Decision Tree
Classification performance (%)
100
75
50
25
0
Balanced Accuracy
Sensitivity
Specificity
Accuracy
AUC
Evaluation metric

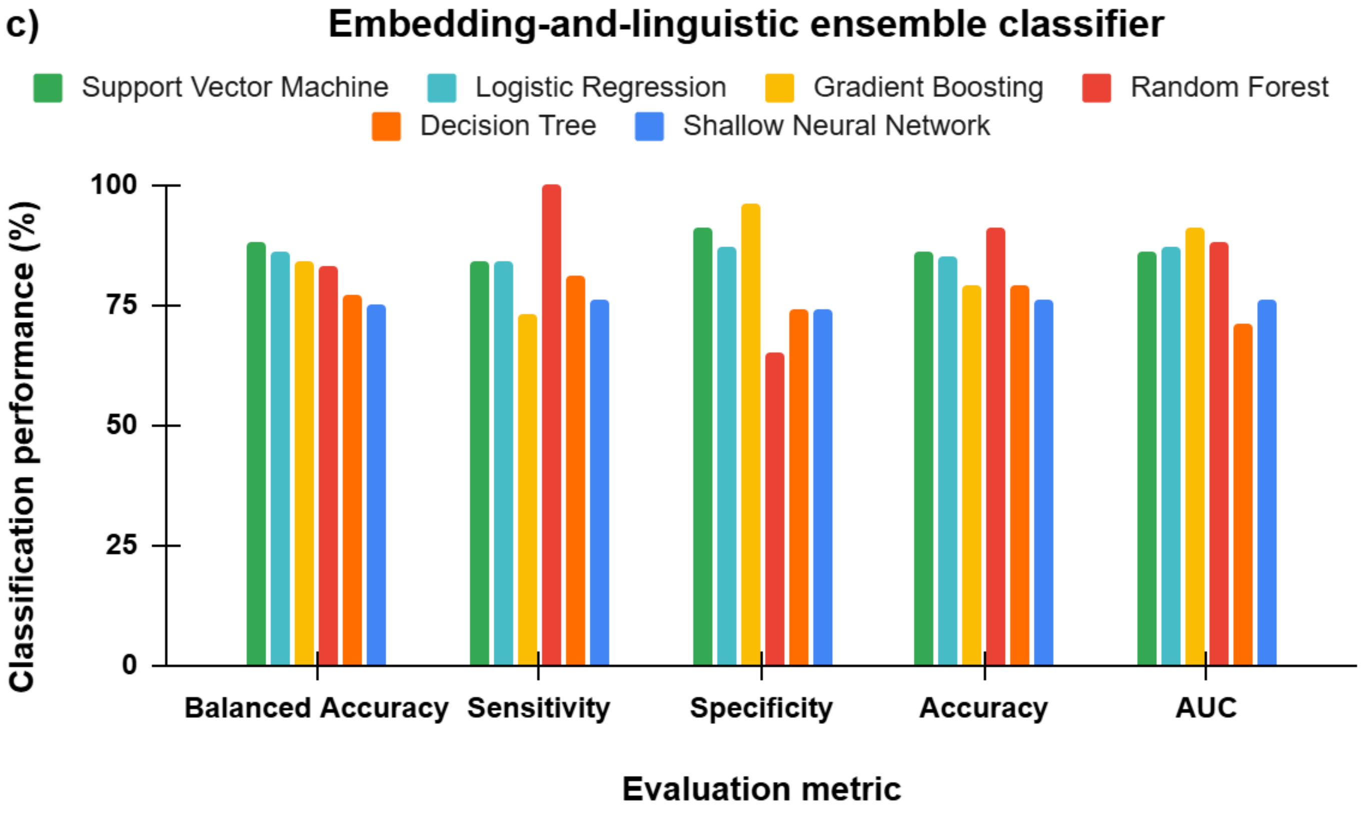
c)
Embedding-and-linguistic ensemble classifier
Support Vector Machine
Logistic Regression
Gradient Boosting
Random Forest
Decision Tree
Shallow Neural Network
Classification performance (%)
100
75
50
25
0
Balanced Accuracy
Sensitivity
Specificity
Accuracy
AUC
Evaluation metric

**Supplementary Figure 5. Balanced accuracy across independent embedding classifiers and algorithms.** Comparison of balanced accuracy (%) achieved for each embedding source (GloVe, BERT, Mistral, OpenAI)[22-25] across candidate classification algorithms. Balanced accuracy represents the average of sensitivity (the proportion of participants with language impairment correctly identified) and specificity (the proportion of participants without language impairment correctly identified). Chance-level performance for balanced accuracy is 50%. As seen in the black bar, balanced accuracy is also shown for each embedding source's best algorithm, defined as the algorithm that achieved the highest balanced accuracy score for the embedding source.

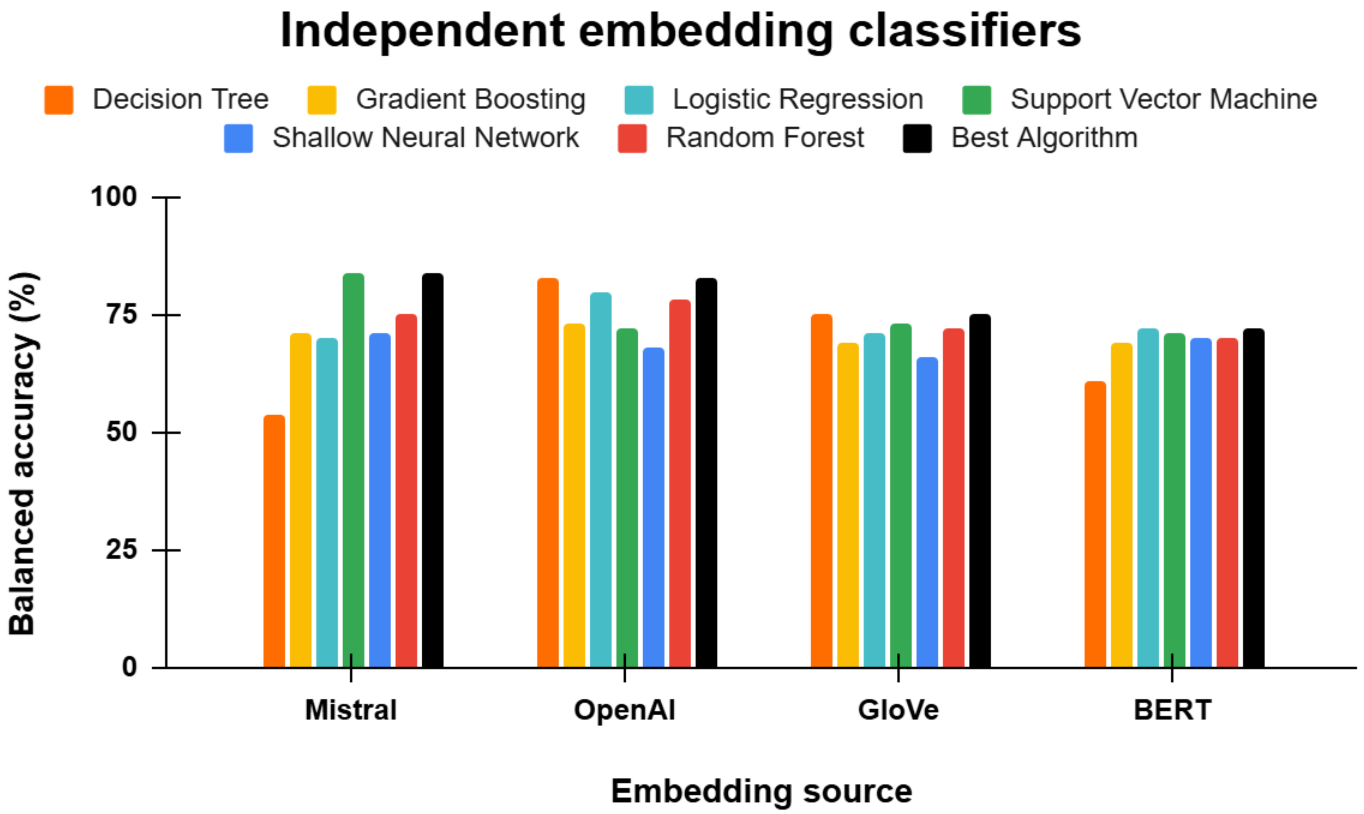

**Supplementary Figure 6. Participant selection and diagnostic classification flowchart.** Flow diagram depicting participant inclusion, exclusion, and diagnostic group assignment. Of 101 acute left-hemisphere stroke participants who completed story retelling, 15 were excluded: Six due to pre-existing conditions affecting cognition (tumor, n = 3; Alzheimer's disease n = 1; substance abuse, n = 2), seven due to insufficient diagnostic data to confirm language status ((picture naming error >10% alongside missing NIHSS Item 9 and speech language pathologist (SLP)evaluations), and 2 due to missing all three diagnostic measures. The final cohort (N = 86) comprised participants classified as having Language Impairment (n = 63), meeting at least one ground-truth criterion; and No Language Impairment (n = 23). SLP = Speech-Language Pathologist; EMR = Electronic Medical Record; NIHSS = National Institutes of Health Stroke Scale.

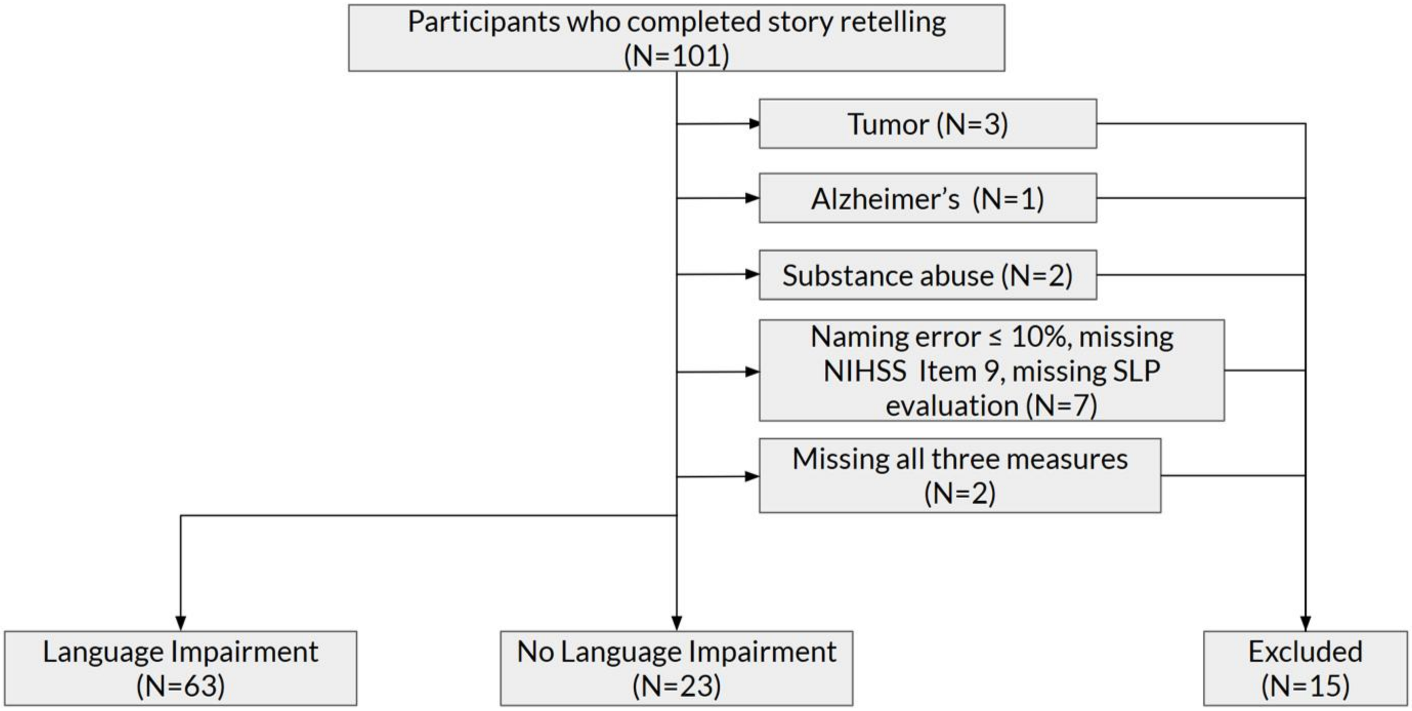

**Supplementary Tables.**

**Supplementary Table 1. Distribution of ground-truth criteria across Language Impairment (N=63) and No Language Impairment (n = 23) groups.**

| **Picture Naming Ability** | **SLP Diagnosis in EMR** | **NIHSS Item 9** | **Frequency (n)** |
|---|---|---|---|
| **Language Impairment (n = 63)** | | | |
| LI | Unavailable | Unavailable | 15 |
| LI | Unavailable | NLI | 14 |
| LI | Unavailable | LI | 2 |
| LI | LI | Unavailable | 3 |
| LI | LI | NLI | 7 |
| LI | LI | LI | 5 |
| LI | NLI | Unavailable | 1 |
| LI | NLI | NLI | 1 |
| NLI | LI | Unavailable | 3 |
| NLI | LI | NLI | 2 |
| NLI | LI | LI | 5 |
| Unavailable | LI | LI | 1 |
| Unavailable | LI | Unavailable | 4 |

| Picture Naming Ability | SLP Diagnosis in EMR | NIHSS Item 9 | Frequency (n) |
|---|---|---|---|
| **No Language Impairment (n = 23)** | | | |
| NLI | Unavailable | NLI | 18 |
| NLI | NLI | Unavailable | 2 |
| NLI | NLI | NLI | 3 |

*Note.* Participants were classified as having language impairment (LI) if at least one of the three diagnostic criteria indicated language impairment. Across the total analyzed cohort (n = 86), missing data frequencies for each criterion were n = 49 for SLP diagnosis in EMR, n = 28 for NIHSS Item 9, and n = 5 for Picture Naming. Within the LI group (n = 63), 5 participants met all three criteria, 18 met two criteria, and 40 met one criterion. Participants were classified as having no language impairment (NLI) if all available diagnostic criteria indicated no impairment (n = 23). SLP = Speech-Language Pathologist; EMR = Electronic Medical Record; NIHSS = National Institutes of Health Stroke Scale.

**Supplementary Table 2. Distribution of source values for patients misclassified by the best performing (embedding-only ensemble) classifier (N=13).**

| True classification | Picture Naming Ability | SLP Diagnosis in EMR | NIHSS item 9 | Frequency (n) |
|---|---|---|---|---|
| LI | LI | Unavailable | Unavailable | 2 |
| LI | LI | Unavailable | NLI | 3 |
| LI | LI | LI | Unavailable | 2 |
| LI | LI | LI | NLI | 2 |
| LI | NLI | LI | Unavailable | 3 |
| LI | NLI | LI | NLI | 1 |

*Note*. All 13 misclassifications were false negatives (true LI classified as NLI); there were zero false positives. LI: Language Impairment; NLI = No Language Impairment; SLP = Speech Language Pathologist; EMR = Electronic Medical Record; NIHSS = National Institute of Health Stroke Scale.

**Supplementary Table 3. Operational definitions and computational derivation of discrete linguistic features.**

| Feature | Definition |
|---|---|
| **Discourse level** | |
| Semantic thematic co-occurrence[46] | The degree to which two consecutive words co-occur. Derived as the cosine similarity between the GloVe[22] embeddings of two consecutive words, averaged across the transcript. |
| Surprisal[15] | The degree to which a word is unexpected given preceding words, calculated for each word, averaged across the transcript. Derived from the large language model Mistral-7B-v0.1[61] with Minicons[48]. |
| Global coherence*[45,49] | The degree to which each utterance relates to the topic under discussion, derived using code from Hoffman et al.[45,49]. |
| **Utterance level** | |
| Mean sentence length | Mean number of words per sentence (total words / total sentences). |
| Moving average type token ratio | No. unique words / no. words, calculated using a moving window of 5 words. |
| **Word level** | |
| No. of words | Total number words in the transcript |
| Proportion pronouns | No. pronouns / (no. nouns + no. pronouns) |
| Proportion verbs | No. verbs / (no. nouns + no. verbs) |
| Proportion closed-class words produced | No. closed-class words / no. words |

| Feature | Definition |
|---|---|
| Semantic diversity[62] | The degree to which the different contexts associated with a given word vary in their meanings, averaged across all open-class words. |
| Age of acquisition[63] | Approximate age when a word is learned, averaged across all open-class words. |
| No. of core lexical items | Number of words produced that belong to a pre-defined set of words commonly used by neurologically healthy controls when retelling Cinderella e.g., “slipper” (core lexicon item[42]). If a core lexical item was produced multiple times, it only counts once towards the total count. |
| Unigram log probability | A measure of word frequency normalized by corpus size, averaged across all open-class words. Derived from the SUBTLEX corpus[64]. |
| Concreteness[65] | The degree to which a concept denoted by a word refers to a perceptible entity, averaged across all open-class words |

*Note*. Features were extracted from automated Whisper transcripts. All features except global coherence were derived using custom SpaCy pipelines adapted from Hilsabeck et al.[28]. Open-class words are defined as nouns, verbs, adjectives, and adverbs. Closed-class words comprise all words excluding open-class words and -ly adverbs.
